# Natural language processing for scalable feature engineering and ultra-high-dimensional confounding adjustment in healthcare database studies

**DOI:** 10.1101/2025.01.30.25321403

**Authors:** Richard Wyss, Jie Yang, Sebastian Schneeweiss, Joseph M. Plasek, Li Zhou, Thomas Deramus, Janick G. Weberpals, Kerry Ngan, Theodore N. Tsacogianis, Kueiyu Joshua Lin

**Author notes:** **Address for Correspondence**: Richard Wyss, PhD, Division of Pharmacoepidemiology and Pharmacoeconomics, Department of Medicine, Brigham and Women’s Hospital, Harvard Medical School 1620 Tremont St. Suite 3030, Boston, MA 02120.

## Abstract

**Background:** To improve confounding control in healthcare database studies, data-driven algorithms may empirically identify and adjust for large numbers of pre-exposure variables that indirectly capture information on unmeasured confounding factors (‘proxy’ confounders). Current approaches for high-dimensional proxy adjustment do not leverage free-text notes from EHRs. Unsupervised natural language processing (NLP) technology can scale to generate large numbers of structured features from unstructured notes.

**Objective:** To assess the impact of supplementing claims data analyses with large numbers of NLP generated features for high-dimensional proxy adjustment.

**Methods:** We linked Medicare claims with EHR data to generate three cohorts comparing different classes of medications on the 6-month risk of cardiovascular outcomes. We used various NLP methods to generate structured features from free-text EHR notes and used LASSO regression to fit several PS models that included different covariate sets as candidate predictors. Covariate sets included features generated from claims data only, and claims data plus NLP-generated EHR features.

**Results:** Including both claims codes and NLP-generated EHR features as candidate predictors improved overall covariate balance with standardized differences being <0.1 for all variables. While overall balance improved, the impact on estimated treatment effects was more nuanced with adjustment for NLP-generated features moving effect estimates further in the expected direction in two of the empirical studies but had no impact on the third study.

**Conclusion:** Supplementing administrative claims with large numbers of NLP-generated features for ultra-high-dimensional proxy confounder adjustment improved overall covariate balance and may provide a modest benefit in terms of capturing confounder information.

## 1. INTRODUCTION

Healthcare data generated from clinical practice, including electronic health records (EHR) and insurance claims, can complement randomized controlled trials to provide evidence on the effects of medical products to support clinical decisions.^1^ However, estimating causal effects from these data sources, so called real-world evidence (RWE), can be challenging due to confounding caused by non-randomized treatment allocation and poorly measured information on comorbidities.^2,3^ Approaches to mitigate confounding bias would ideally be based on causal diagrams and expert knowledge for variable selection.^4^ Covariate adjustment based on expert knowledge alone, however, is not always adequate because some confounders may not be considered by researchers or not be directly measurable in such secondary healthcare data.

To improve confounding control in RWE studies, data-driven algorithms can be used to empirically identify and adjust for large numbers of pre-exposure variables that indirectly capture information on unmeasured or unspecified confounding factors (‘proxy’ confounders).^5–8^ A growing literature has shown that supplementing investigator-specified variables with large numbers of empirically identified features can often improve confounding control compared to adjustment based on investigator-specified variables alone.^5–13^ Current approaches for high-dimensional proxy adjustment, however, require data to be in a structured format (e.g., claims and structured EHR data), leaving unstructured EHR text information underutilized for confounding control. Leveraging this information can be challenging since patient-reported records are often recorded in free-text documents that are not readily analyzable at a large scale.

Recent work has demonstrated that unsupervised natural language processing (NLP) technology can scale to generate large numbers of structured features from unstructured clinical documents.^14,15^ However, the added value of supplementing administrative claims data with large numbers of NLP-generated EHR features to improve high-dimensional proxy confounding control in healthcare database studies remains unclear. Here, we use three empirical studies to investigate the added value of supplementing administrative claims with high-dimensional sets of NLP-generated features from time-indexed EHR notes for causal analyses. We consider several NLP methods for generating structured features from pre-exposure free-text clinical notes and observe how adjustment for different covariate sets impacts covariate balance and effect estimates after PS weighting. Our objective is to assess if unsupervised NLP tools can leverage information in EHR free-text notes to supplement claims data for improved high-dimensional proxy adjustment in healthcare database studies.

## 2. METHODS

Figure 1 summarizes the analytic pipeline we used for feature generation and high-dimensional proxy adjustment. Each step is described in detail below.

**Figure 1.**
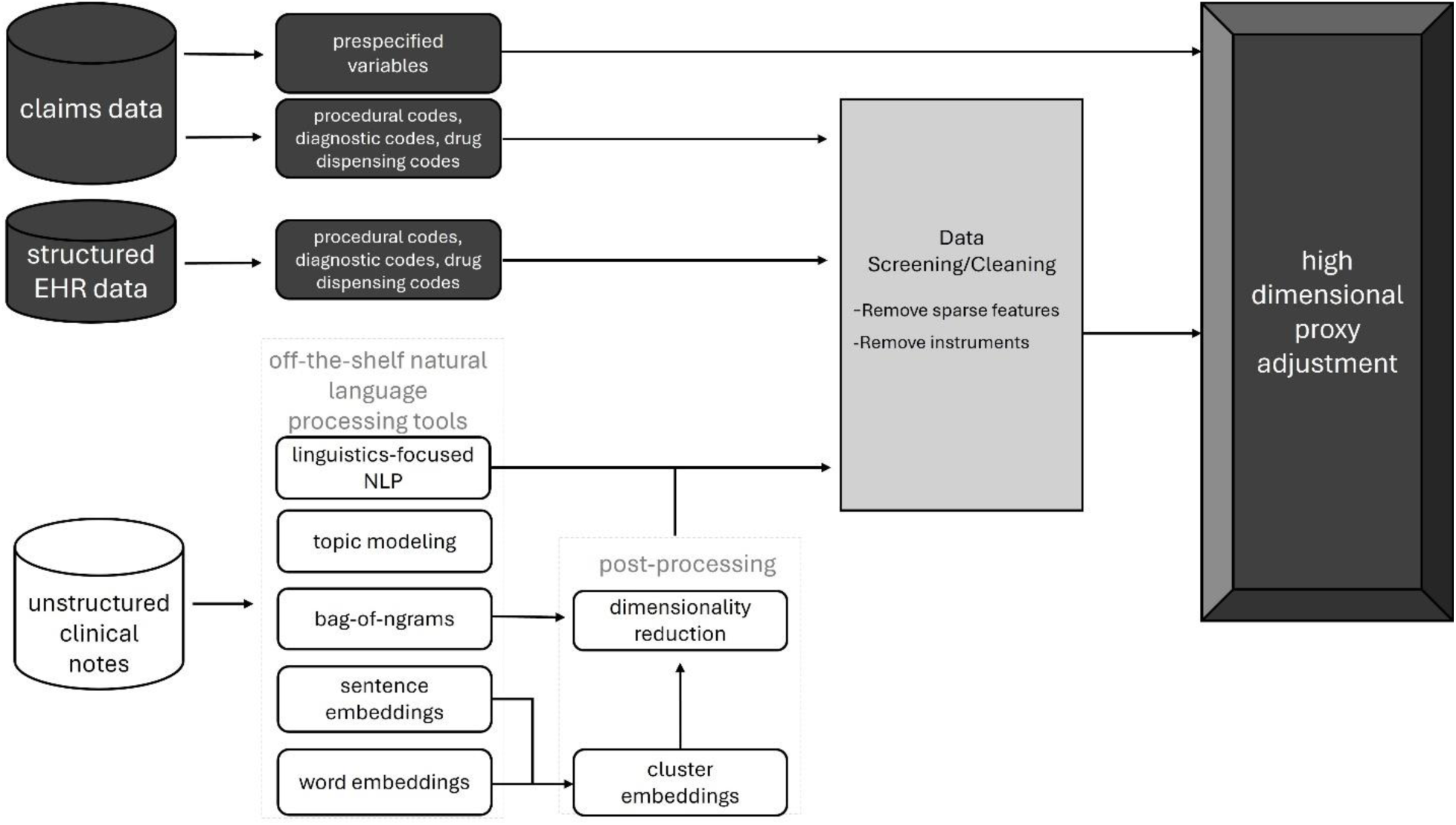
Analytic pipeline illustrating the process for feature engineering of structured and unstructured data for high-dimensional proxy confounder adjustment.

**Figure 2.**
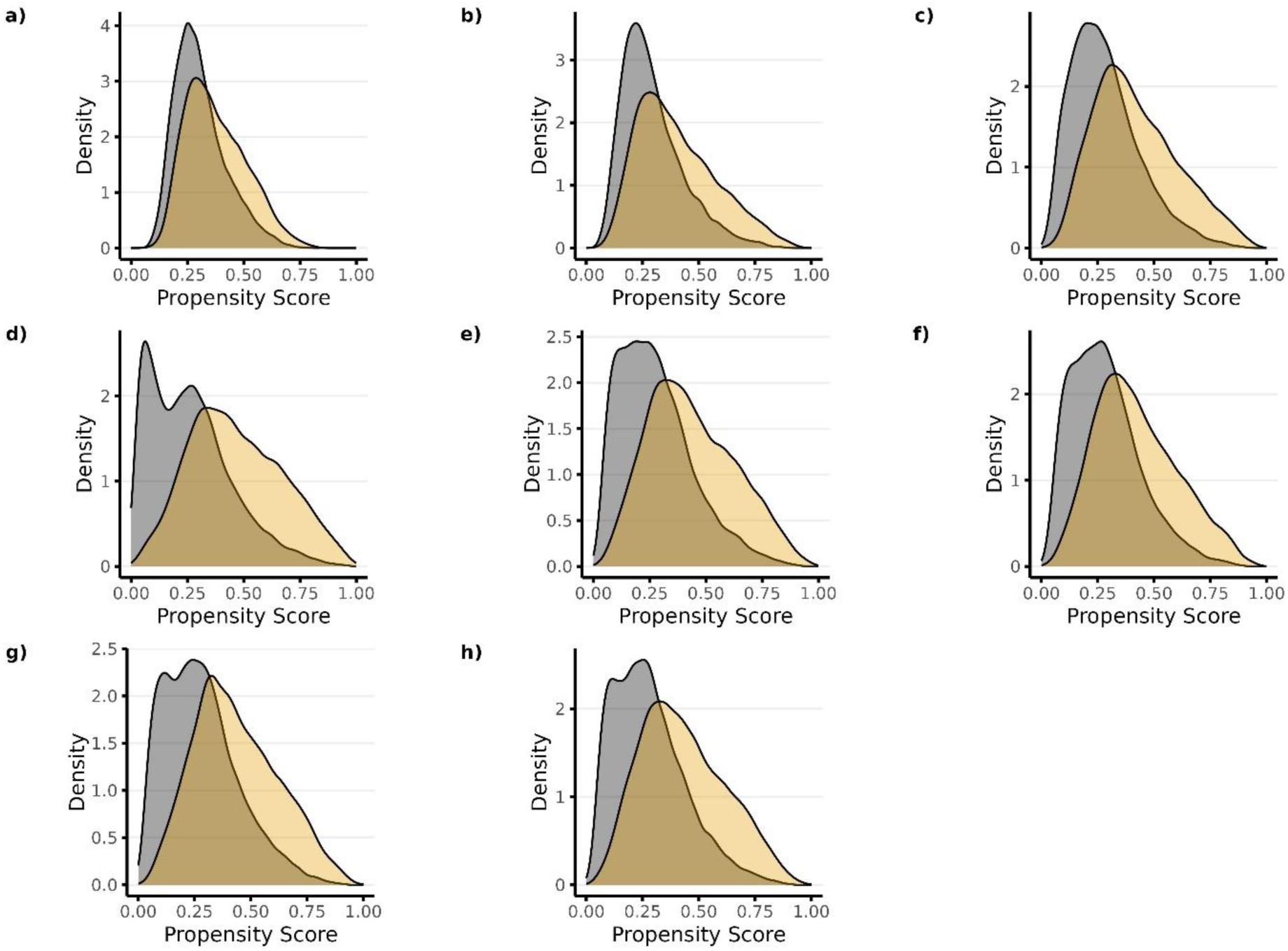
Propensity score distributions plotted across treatment groups for each propensity score model in Study 1 (high-dose PPI vs low-dose PPI). Panels a) through h) correspond to PS models 1-8, respectively. Candidate predictors for each model included the following: Model 1—researcher-specified variables only; Model 2— researcher-specified variables + claims codes; Model 3—research-specified variables + claims codes + EHR codes; Model 4—research specified variables + claims codes + EHR codes + NLP-generated features using ngrams; Model 5—researcher-specified variable + claims codes + EHR codes + NLP-generated features using mterms; Model 6— researcher-specified variable + claims codes + EHR codes + NLP-generated features using sentence embeddings; Model 7—researcher-specified variable + claims codes + EHR codes + NLP-generated features using word embeddings with bert; Model 8—researcher-specified variables + claims codes + EHR codes + NLP-generated features using word embeddings with glove.

### 2.1. Data Source

We linked longitudinal claims data from the US Medicare system to the Research Patient Data Registry (RPDR) from 2007/01/01 to 2017/12/31. The RPDR data repository is based on all inpatient and outpatient activities of the Mass General Brigham (MGB), the largest healthcare delivery network in the greater Boston area including 2 academic medical centers, 14 community hospital and more than 100 satellite clinics. RPRD data records all medical records electronically, including diagnoses, procedures, test results (lab tests, imaging, biopsies, etc.), prescribing, and free text notes for all inpatient and outpatient services. Linking Medicare claims with the RPDR data repository helped reduce data leakage due to EHR-discontinuity (i.e., missing information from medical encounters provided outside the MGB network).^16^

### 2.2. Study population

Based on Medicare fee-for-service beneficiaries aged 65 years or older, we generated 3 cohorts:

1. *HTN Cohort*: A cohort of patients with hypertension comparing angiotensin-converting enzyme inhibitors (ACEi) vs beta blockers on the risk of hyperkalemia.
2. *Analgesics Cohort*: A cohort of patients using analgesics comparing NSAIDs vs Opioids in patients with a history of osteoarthritis (OA) in terms of risk of acute kidney injury (AKI).
3. *PPI Cohort*: A cohort of patients using proton pump inhibitors (PPIs) comparing high-vs low-dose PPI use in patients with a history of peptic ulcer in terms of risk of gastrointestinal (GI) bleeding.

For each empirical cohort, we identified individuals who initiated the treatment (or comparator) after no use of either the treatment or comparator medication in the previous year (new-user design).^17,18^ The cohort entry date is the first record date of the medication use. To ensure that the study population had adequate information recorded in our data source, we required the study population to have at least 364 days of Medicare continuous enrolment in parts A (inpatient coverage), B (outpatient coverage), and D (prescription coverage). Characteristics for each study cohort are shown in Table 1. Further details of each generated cohort and study design are provided in the Supplemental Appendix.

**Table 1.**
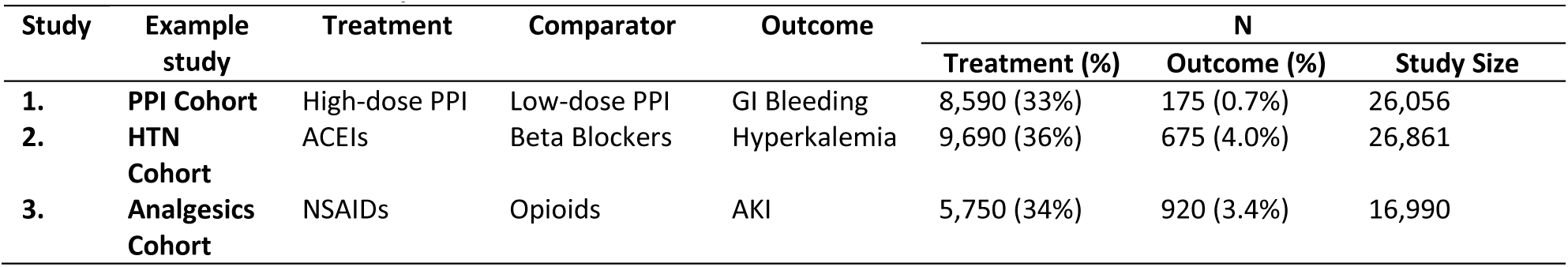
Characteristics of study cohorts.

These studies were chosen because they 1) present challenges in terms of confounding due to selective prescribing (confounding by indication); and 2) address clinically relevant questions. For example, while RCTs suggest no association between PPI dose and GI bleeding complications,^19,20^ our unadjusted analysis showed that high-dose PPI was associated with a higher risk of GI bleeding when compared to low-dose PPI. This is likely due to confounding where individuals who have a history of more severe gastrointestinal bleeding were channeled to higher doses of PPI. For the HTN Cohort, it is common for patients at high risk of hyperkalemia to be channeled away from ACE inhibitors. As a result, we observed an apparent protective effect against hyperkalemia associated with ACE inhibitors when compared with beta-blockers, which is likely confounded.^21^ We would expect effective confounding adjustment would move the estimates toward the null.^22^ Finally, for the Analgesics Cohort, previous studies suggest that NSAIDs are nephrotoxic and can cause AKI.^23,24^ Consequently, patients with a history of kidney diseases are more likely to be prescribed an opioid potentially leading to strong confounding by indication showing an apparent decreased risk of AKI associated with NSAIDs when compared to opioids. This confounded association was observed in our unadjusted analysis and an effective confounding adjustment would be expected to move the estimates toward the harmful effects associated with NSAIDs.

### 2.3. Generating Structured Features from Unstructured EHR Free Text Notes

We applied 5 different unsupervised NLP approaches ranging from basic statistical features to contextual deep-learning representation features, to generate structured features from unstructured free-text notes. For all approaches, we considered unstructured information during the 365 days before cohort entry. Additional details and code for NLP generation are available on GitHub:^25^

- *bag-of-words or bag-of-n-grams*: An n-gram is a sequence of consecutive items (in this case, words), where “unigram” refers to a single word, “bigram” refers to 2 consecutive words, and so on. Each document was tokenized and processed into unigrams and bigrams. We excluded stop words, i.e., words that occur frequently but convey little semantic meaning, such as articles (e.g., “a”, “the”) and prepositions (e.g., “in”, “on”).
- *MTERMS*: An automatic tool that extracts clinical terms or concepts from EHRs. Using lexical and term pattern matching methods, MTERMS can extract medical problems, medications, adverse reactions, and allergies.^26^
- *Word Embeddings (GloVe)\**: Word embeddings are commonly used to convert each word into a continuous vector that can capture the semantic similarity between words. With word embeddings, if two words have similar neighboring words, then their embedding vectors will be close to each other, reflecting their similarity, even if the two words do not share morphological similarity.^27^
- *Contextual Word Embeddings (BioBERT)*: An advanced contextual word embeddings model that takes the context of a word into consideration when generating the embedding. Compared to GloVe word embeddings, contextual word embeddings can provide unique embeddings for the same word under different contexts, ensuring fine-grained semantic information for the word. The BioBERT model was trained with biomedical text, which makes it specifically tuned for tasks in the biomedical domain, enhancing its ability to understand and process medical terminology more accurately.^28^
- *Sentence Embeddings (BERT):* Similar to contextual word embeddings, sentence embeddings with BERT use contextual representation but represent the entire sentence rather than individual words. It takes the sentence as the basic unit to generate candidate features. For the two BERT approaches we used the KMeans algorithm (MiniBatchKMeans) to cluster all the embeddings and represent patients with a binary cluster feature.^29^

It is important to emphasize that a key factor for the scalability of using NLP tools to generate large numbers of structured features for real-time clinical decision support is that researchers can remain agnostic to the format and content of the processed information. The NLP methods described above were used to automatically identify concepts or patterns from EHR free-text notes which were then fed into a LASSO regression to model the treatment choice as a propensity score for high-dimensional proxy adjustment. We were not concerned about the specific clinical meaning of the extracted features but only their potential for confounding the specified causal analyses. In other words, the application of NLP tools in this setting does not require time-intensive training data creation and reference-standard annotation through manual chart review.

### 2.4. Data Screening, Propensity Score Development, and Confounding Adjustment

#### 2.4.1. Data screening

For each empirical study, baseline covariates (features) included several dozen researcher-specified variables (which included demographic variables like age and sex), thousands of additional claims codes, and thousands of NLP-generated features from EHR. The researcher-specified variables were constructed using claims data only that were determined based on diagnostic and procedural codes along with all NDC drug codes. NLP-generated features only included EHR information in the year before cohort entry. We then screened for and excluded from the analysis claims codes and NLP-generated features with a prevalence < 0.01.

We then screened features that were strongly associated with treatment choice but not or only weakly with the outcome and thus behaved like instrumental variables (IVs). These were excluded since adjusting for IVs harms the properties of causal effect estimates.^30–32^ We ranked each variable based on its marginal correlation with treatment and manually examined the top-ranked variables. Those of the top-ranked variables that were not deemed to be risk factors for the outcome based on clinical domain knowledge were excluded from the analysis. While not comprehensive, we were able to examine the strongest predictors of treatment to exclude instruments that are most likely to be harmful to causal analyses (Supplemental Appendix).

#### 2.4.2. Propensity score models

We used Lasso regression to fit 8 different propensity score models, each containing one of the following covariate sets as candidate predictors:

1) Researcher specified variables only,
2) Researcher specified variables + automated feature extraction from claims codes,
3) Researcher specified variables + automated feature extraction from claims codes + EHR codes
4) Researcher-specified variables + automated feature extraction from claims codes + EHR codes + NLP features generated using Ngrams.
5) Researcher-specified variables + automated feature extraction from claims codes + EHR codes + NLP features generated using MTERMS.
6) Researcher-specified variables + automated feature extraction from claims codes + EHR codes + NLP features generated using sentence embeddings (BERT).
7) Researcher-specified variables + automated feature extraction from claims codes + EHR codes + NLP features generated using word embeddings (BioBERT).
7) Researcher-specified variables + automated feature extraction from claims codes + EHR codes + NLP features generated using word embeddings (GloVe).

To avoid overfitting the Lasso PS models we randomly split the data into 10 equally sized non-overlapping groups, trained the Lasso model in 9 of the groups, which was then applied to the 10th group to assign predicted probabilities. This process was repeated 10 times for each model. Theory and simulations have shown that the use of cross-fitting reduces the impact of modeling spurious associations in the data when using data-adaptive algorithms for estimating the PS for causal inference.

We evaluated both the discrimination and calibration of each model by calculating the C-statistic (AUC) and negative log-likelihood (NLL) using 10-fold cross-validation. Note that high values for the C-statistic correspond with stronger discrimination while lower values for negative log-likelihood correspond with better calibration (more accurate predictions).

#### 2.4.3. Propensity score analyses

We adjusted for PSs using the following weighting approaches:^33–35^

● Inverse Probability Treatment Weights 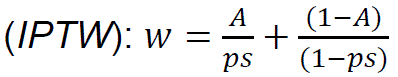
● *Overlap Weights*: 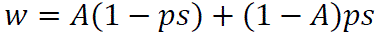
● *Matching Weights*: 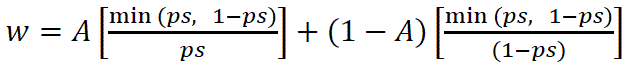

With *A* being the binary treatment choice and *ps* the estimated propensity score.

For each example study, we used a weighted Cox regression to estimate the hazard ratio and 95% confidence intervals over a 6-month follow-up window with an intent-to-treat analysis. We assessed the covariate balance between treatment groups by calculating the standardized differences after PS weighting. For each PS weighted analysis, balance was assessed on variables within the covariate set that was available to be included as candidate predictors in the PS model, as well as all variables across all covariate sets even though they were not eligible to be included in a given model. The latter was done to also assess the balance achieved in NLP-derived features not included in the adjustment. Covariates with standardized differences < 0.1 were assumed to provide adequate confounding control.^36^

## 3. RESULTS

All results for PS analyses shown here used matching weights for covariate adjustment, results using other weights are shown in the Supplemental Appendix.

### 3.1. Predicting Treatment Choice and PS Overlap

The number of candidate predictors within each covariate set, the number of candidate predictors selected by each of the 8 LASSO PS model, and their prediction diagnostics are shown in **Table 2**.

**Table 2.** Prediction diagnostics and hazard ratio estimates when using matching weights for covariate adjustment.

| Example study | Model<sup>a</sup> | # Candidate Predictors | # Predictors Selected | AUC | NLL | HR | 95% CI |
| --- | --- | --- | --- | --- | --- | --- | --- |
| **PPI Cohort** |  |  |  |  |  |  |  |
|  | Unadjusted | ----- | ----- | ----- | ----- | 2.77 | 2.05, 3.74 |
|  | Model 1 | 84 | 84 | 0.66 | 0.60 | 1.80 | 1.31, 2.48 |
|  | Model 2 | 2,152 | 489 | 0.69 | 0.58 | 1.40 | 1.01, 1.95 |
|  | Model 3 | 3,060 | 523 | 0.72 | 0.57 | 1.43 | 1.03, 2.01 |
|  | Model 4 | 35,634 | 806 | 0.77 | 0.52 | 1.38 | 0.95, 1.97 |
|  | Model 5 | 6,870 | 554 | 0.74 | 0.55 | 1.43 | 1.02, 2.01 |
|  | Model 6 | 12,682 | 637 | 0.72 | 0.56 | 1.46 | 1.04, 2.05 |
|  | Model 7 | 13,022 | 809 | 0.74 | 0.55 | 1.40 | 1.00, 1.97 |
|  | Model 8 | 6,051 | 610 | 0.73 | 0.55 | 1.54 | 1.08, 2.16 |
| **HTN Cohort** |  |  |  |  |  |  |  |
|  | Unadjusted | ----- | ----- | ----- | ----- | 0.59 | 0.52, 0.69 |
|  | Model 1 | 82 | 82 | 0.80 | 0.51 | 0.93 | 0.79, 1.11 |
|  | Model 2 | 1,622 | 426 | 0.83 | 0.49 | 0.96 | 0.80, 1.15 |
|  | Model 3 | 2,278 | 535 | 0.86 | 0.45 | 0.99 | 0.83, 1.20 |
|  | Model 4 | 32,424 | 644 | 0.89 | 0.39 | 0.96 | 0.79, 1.16 |
|  | Model 5 | 5,475 | 663 | 0.89 | 0.39 | 1.01 | 0.83, 1.23 |
|  | Model 6 | 11,376 | 969 | 0.86 | 0.45 | 0.96 | 0.79, 1.16 |
|  | Model 7 | 12,176 | 847 | 0.88 | 0.42 | 0.99 | 0.82, 1.20 |
|  | Model 8 | 5,304 | 599 | 0.88 | 0.41 | 0.97 | 0.79, 1.17 |
| **Analgesic Cohort** |  |  |  |  |  |  |  |
|  | Unadjusted | ----- | ----- | ----- | ----- | 0.41 | 0.34, 0.50 |
|  | Model 1 | 77 | 77 | 0.78 | 0.52 | 0.76 | 0.61, 0.95 |
|  | Model 2 | 1,545 | 462 | 0.82 | 0.49 | 0.76 | 0.61, 0.96 |
|  | Model 3 | 2,029 | 454 | 0.84 | 0.46 | 0.79 | 0.63, 0.98 |
|  | Model 4 | 29,394 | 746 | 0.85 | 0.45 | 0.81 | 0.64, 1.02 |
|  | Model 5 | 4,905 | 512 | 0.85 | 0.45 | 0.78 | 0.62, 0.99 |
|  | Model 6 | 10,822 | 432 | 0.84 | 0.47 | 0.79 | 0.63, 0.99 |
|  | Model 7 | 11,881 | 1035 | 0.90 | 0.37 | 0.79 | 0.61, 1.03 |
|  | Model 8 | 4,937 | 655 | 0.84 | 0.46 | 0.80 | 0.64, 1.01 |

<sup>a</sup>**Model 1)** Researcher specified variables only, **Model 2)** Researcher specified variables + automated feature extraction from claims codes, **Model 3)** Researcher specified variables + automated feature extraction from claims codes + EHR codes, **Model 4)** Researcher-specified variables + automated feature extraction from claims codes + EHR codes + NLP features generated using Ngrams, **Model 5)** Researcher-specified variables + automated feature extraction from claims codes + EHR codes + NLP features generated using MTERMS, **Model 6)** Researcher-specified variables + automated feature extraction from claims codes + EHR codes + NLP features generated using sentence embeddings (BERT), **Model 7)** Researcher-specified variables + automated feature extraction from claims codes + EHR codes + NLP features generated using word embeddings (BioBERT), **Model 8)** Researcher-specified variables + automated feature extraction from claims codes + EHR codes + NLP features generated using word embeddings (GloVe).

Prediction diagnostics show that PS models that only included researcher-specified variables resulted in the poorest predictive performance in terms of the C-statistic (AUC) and NLL. Supplementing researcher-specified variables with large numbers of features from claims and NLP-generated EHR features improved predictive performance. Overall, PS models that included Covariate Set 4 as candidate predictors (researcher-specified variables + claims codes + NLP-generated features using ngrams) resulted in the best predictive performance for both the AUC and NLL in comparison with the other covariate sets (**Table 2**).

The improved predictive performance when supplementing researcher-specified variables with large numbers of claims codes and NLP-generated features is reflected in the propensity score distributions plotted across treatment groups (Figure 1 **and Supplemental Figures S1 an S2**). As predictive performance improved, separation in the distribution of the propensity score across treatment increased. PS Model 4, which included Covariate Set 4 as candidate predictors, had the strongest predictive performance and resulted in the largest separation across treatment groups for each of the 3 studies (Plot d in Figure 1 **and Supplemental Figures S1 an S2**). As previous studies have shown, however, PS models with better discrimination (C-statistic) does not necessarily correspond with better confounding control.^37,38^

### 3.2. Covariate Balance

Figures 3-5 show the absolute standardized differences of each covariate across treatment groups for Studies 1-3, respectively, before and after matching weights were used for covariate adjustment. Orange dots represent variables that were not available as candidate predictors for the given PS model. For example, the black dots in Panel a) represent the candidate predictors available for Model 1 which was Covariate Set 1 (in this case only researcher-specified variables), while the orange dots in Panel a) represent all other variables.

**Figure 3.**
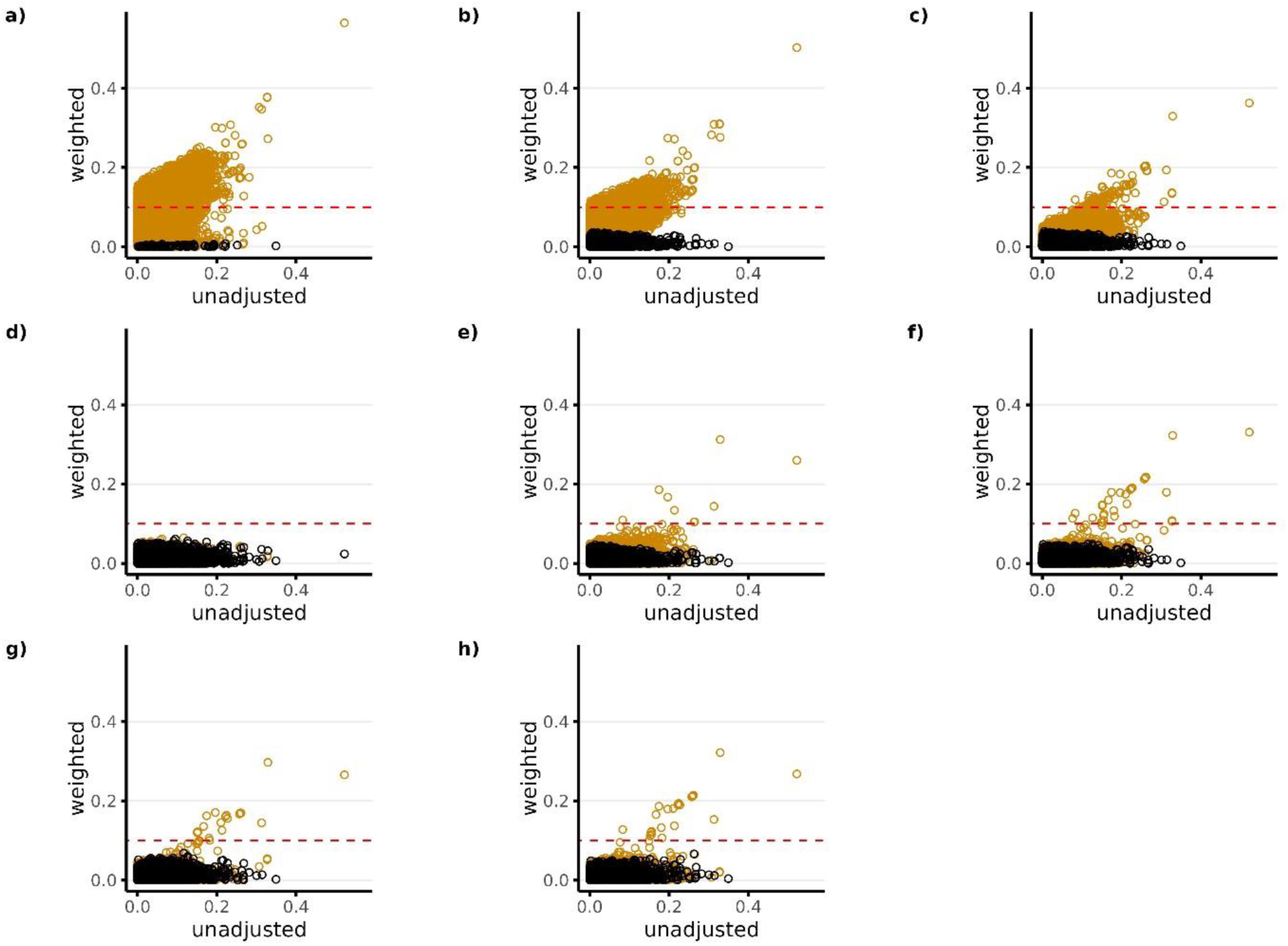
Standardized covariate difference across treatment groups before and after PS weighting (matching weights) for Study 1 (High-dose PPI vs Low-dose PPI). Panels a) through h) correspond to PS models 1-8, respectively. Black dots in each panel represent the candidate predictors that were available for the given PS model. Orange dots represent all other variables that were not available as candidate predictors for the given PS model. The candidate predictors available for each model (black dots) include the following: Model 1—researcher-specified variables only; Model 2—researcher-specified variables + claims codes; Model 3—research-specified variables + claims codes + EHR codes; Model 4—research specified variables + claims codes + EHR codes + NLP-generated features using ngrams; Model 5—researcher-specified variable + claims codes + EHR codes + NLP-generated features using mterms; Model 6—researcher-specified variable + claims codes + EHR codes + NLP-generated features using sentence embeddings; Model 7—researcher-specified variable + claims codes + EHR codes + NLP-generated features using word embeddings with bert; Model 8—researcher-specified variables + claims codes + EHR codes + NLP-generated features using word embeddings with glove. The red horizontal dotted line represents the largest level of balance after PS weighting that was considered adequate (<0.1).

**Figure 4.**
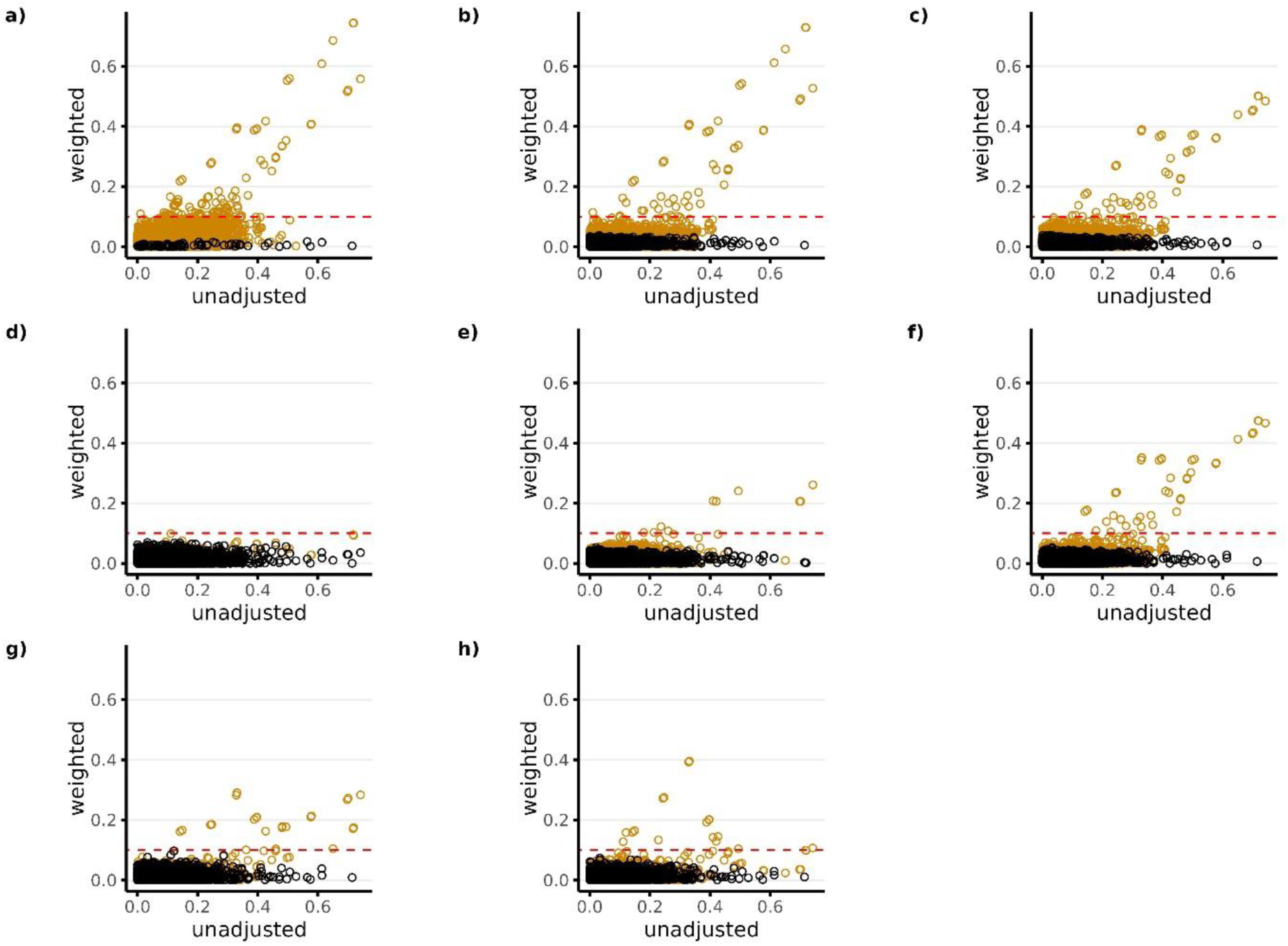
Standardized covariate difference across treatment groups before and after PS weighting (matching weights) for Study 2 (ACEIs vs Beta Blockers). Panels a) through h) correspond to PS models 1-8, respectively. Black dots in each panel represent the candidate predictors that were available for the given PS model. Orange dots represent all other variables that were not available as candidate predictors for the given PS model. The candidate predictors available for each model (black dots) include the following: Model 1—researcher-specified variables only; Model 2—researcher-specified variables + claims codes; Model 3—research-specified variables + claims codes + EHR codes; Model 4—research specified variables + claims codes + EHR codes + NLP-generated features using ngrams; Model 5—researcher-specified variable + claims codes + EHR codes + NLP-generated features using mterms; Model 6—researcher-specified variable + claims codes + EHR codes + NLP-generated features using sentence embeddings; Model 7—researcher-specified variable + claims codes + EHR codes + NLP-generated features using word embeddings with bert; Model 8—researcher-specified variables + claims codes + EHR codes + NLP-generated features using word embeddings with glove. The red horizontal dotted line represents the largest level of balance after PS weighting that was considered adequate (<0.1).

**Figure 5.**
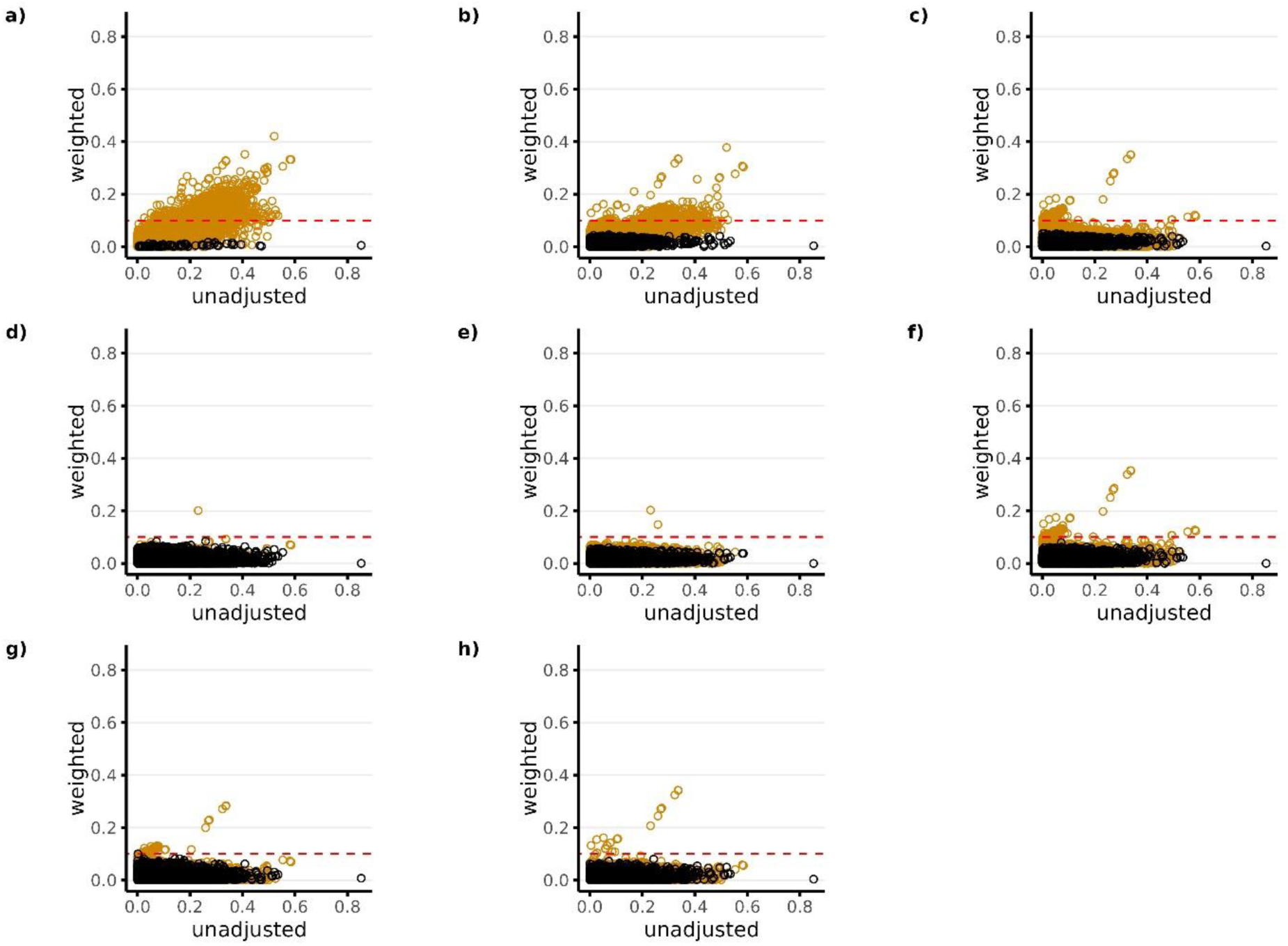
Standardized covariate difference across treatment groups before and after PS weighting (matching weights) for Study 3 (NSAIDs vs Opioids). Panels a) through h) correspond to PS models 1-8, respectively. Black dots in each panel represent the candidate predictors that were available for the given PS model. Orange dots represent all other variables that were not available as candidate predictors for the given PS model. The candidate predictors available for each model (black dots) include the following: Model 1—researcher-specified variables only; Model 2—researcher-specified variables + claims codes; Model 3—research-specified variables + claims codes + EHR codes; Model 4—research specified variables + claims codes + EHR codes + NLP-generated features using ngrams; Model 5—researcher-specified variable + claims codes + EHR codes + NLP-generated features using mterms; Model 6—researcher-specified variable + claims codes + EHR codes + NLP-generated features using sentence embeddings; Model 7—researcher-specified variable + claims codes + EHR codes + NLP-generated features using word embeddings with bert; Model 8—researcher-specified variables + claims codes + EHR codes + NLP-generated features using word embeddings with glove. The red horizontal dotted line represents the largest level of balance after PS weighting that was considered adequate (<0.1).

All PS models performed well in terms of balancing the covariates that were available as candidate predictors for the given model (black dots), with absolute standardized differences being <0.1 for all models across all studies. However, in terms of balancing covariates that were not available as candidate predictors for the given model (orange dots), results show that the models that did not include NLP-generated EHR features as candidate predictors in the LASSO PS models (PS Models 1-3) resulted in large imbalances in the NLP features after PS weighting. This is reflected in Plots a) through c) in Figures 3-5 with a large proportion of the orange dots having standardized differences >0.1. Among the models that included NLP-generated features (PS Models 4-8), Model 4 performed best across all studies in terms of balancing covariates that were not available as candidate predictors (orange dots) with absolute standardized differences being <0.1 for all covariates in studies 1 and 2, and <0.1 for all but 1 covariate in study 3. Overall patterns for covariate balance were similar when using overlap weights for covariate adjustment (**Supplemental Figures S3-S5**). However, when using inverse probability weighting, we found that it was difficult to consistently balance covariates for the majority of models across all studies, regardless of whether or not they were available as candidate predictors (**Supplemental Figures S6-S8**).

### 3.3. Treatment Effect Estimates

**Table 2** also shows the estimated hazard ratios and 95% confidence intervals after using PS matching weights for covariate adjustment. The estimated treatment effects after PS-weighting moved the hazard ratios in the expected direction for each study based on previous trial and clinical evidence discussed above. For example, trial evidence suggests that there is a null relationship between PPI dose and GI bleeding (expected HR close to 1). The unadjusted estimate comparing hi-vs. low-dose PPI was strongly impacted by confounding by indication with the hazard ratio being 2.77 (2.05, 3.74). Adjustment for each covariate set resulted in substantial movement in the estimated treatment effect towards the null with adjustment for Covariate Set 4 (PS Model 4) resulting in an estimated effect with the 95% CI including the null (HR: 1.38; 95% CI: 0.95, 1.97).

For the HTN cohort comparing ACE inhibitors versus beta-blockers on the risk of acute kidney injury, effect estimates were impacted by strong confounding by indication with the unadjusted estimate showing a protective effect for individuals initiating ACE inhibitors with a HR of 0.59 (0.52, 0.69). Adjustment for each covariate set moved the estimated effect towards the null with the 95% CI of all 8 models including the null. Adjustment for Covariate Set 5 (PS Model 5) resulted in an estimated effect that was furthest from the unadjusted estimate with a HR of 1.01 (0.83, 1.23).

For the Analgesics cohort comparing the effect of opioids vs. NSAIDs on the risk of risk of renal failure, results in **Table 2** show strong confounding by indication with the unadjusted estimate showing a protective effect for individuals initiating NSAIDs with a HR of 0.41 (0.34, 0.50). Similar to the PPI study, adjustment for each covariate set moved the estimated effect towards the null with adjustment for Covariate Set 4 resulting in an estimated effect that was furthest from the unadjusted estimate with a HR of 0.81 (0.64, 1.02).

## 4. DISCUSSION

We investigated the impact of supplementing administrative claims with NLP-generated features from EHR free-text notes when using various propensity score-based weighting methods for high-dimensional proxy adjustment. After fitting LASSO PS models, we found that supplementing prespecified variables and claims codes with additional NLP-generated features using n-grams performed best in terms of treatment prediction. Interestingly, the overall covariate balance in all 3 empirical studies also improved and moved treatment effect estimates furthest from the unadjusted estimates in a direction consistent with prior expectation in 2 of the 3 studies. In our experiments, NLP tools more sophisticated than n-grams, such as MTERMS, and word or sentence embeddings, did not further improve covariate balance or confounding adjustment.

The use of NLP technology to leverage information from unstructured free-text EHR notes to improve confounding adjustment in healthcare database studies is a growing area of work.^39–43^ To our knowledge, this study is the first to apply and compare the scalability and performance of several alternative NLP tools for purposes of high-dimensional proxy adjustment. These findings indicate that large-scale NLP-enabled feature generation is quite feasible and may provide additional confounder information when conducting high-dimensional proxy adjustment. This may result in improved effect estimates, although differences in effect estimates were modest in our example studies, and it supports automated approaches to confounding control.

When controlling for a large number of variables, we found that weighting methods that down-weight the tails of the PS distribution (matching weights and overlap weights) were more robust in balancing covariates compared to inverse probability weighting. This finding was consistent across all 3 empirical studies and is also consistent with previous studies that have compared alternative weighting approaches.^44^ Because people with extreme PS values represent those for whom the confounding may be refractory, PS weighting approaches that down-weight the tails of the PS distribution may also mitigate unmeasured confounding that is likely stronger in the tails of the PS distribution.^45^ However, the tradeoff is that the target population for matching weights and overlap weights cannot be defined a priori and is dependent on the distribution of the estimated PS. This creates additional challenges when comparing effect estimates across different PS models as it can be unclear if movement in the estimated treatment effect is due to better confounding control or heterogeneity in the treatment effect caused by changing target populations. Ideally, we would target the same population across all analyses by weighting to the same population (e.g., using inverse probability weights); however, across all 3 empirical studies we were only able to adequately balance covariates when using methods that down-weight the tails of the PS when controlling for high-dimensional sets of variables. Consequently, we did not focus on results from analyses using inverse probability weights.

Some additional limitations deserve attention. First, other factors including random chance and hidden biases not captured through PS weighting could also contribute to unpredictable movement in estimated effects and a true ‘gold standard’ is never known with certainty. Consequently, as discussed previously, methodological comparisons of estimated treatment effects in real-world data examples are inherently limited. However, such comparisons are widely used and can be informative, particularly when consistent trends are observed across multiple studies increasing confidence in the robustness of findings.

Second, while we explored several NLP methods for generating structured features from free-text notes, there are many NLP tools that were not considered or customized to our EHR data due to resource constraints. Future work could explore additional NLP tools such as large language models, for extracting confounder information from EHR data, as well as training embeddings on our local EHR data for better representing features in EHRs. In addition, different ways of modeling the NLP-generated features may also improve the performance. For example, in this study we simply created dichotomous variables from the n-gram and other NLP output. Other options could consider modeling the NLP output as categorical or continuous features, such as using the term frequency and the reciprocal document frequency (tf-idf) of the n-gram output or the weights of embedding clusters. These approaches help integrate the fine-grained contribution weights of NLP-generated features. In this study, we chose to focus on generating binary features from the NLP output to simplify the functional form of the PS model and reduce the likelihood of model misspecification. While more flexible nonparametric models could potentially be used to model complex relationships between continuous/categorical features and treatment, both theory and simulations have shown that using flexible nonparametric models for PS estimation comes at a cost of slow convergence rates, which can harm the properties of causal estimators, particularly in high-dimensional models.^46–48^ A thorough comparison of modeling more complex NLP-output with flexible nonparametric models is beyond our scope.

In conclusion, we found that off-the-shelf NLP tools can scale to generate large numbers of structured features from free-text EHR notes. We found that supplementing administrative claims with large numbers of NLP-generated EHR features improved overall covariate balance in PS-weighted analyses, but the observed impact on estimated treatment effects was incremental and more complex NLP tools had no advantage over simple n-grams.

## Author Approval

All authors of this paper have read and approved the final version submitted.

## Data Availability

Data used in the present study are not publicly available due to data use agreements.

